# Household secondary attack rate of COVID-19 by household size and index case characteristics

**DOI:** 10.1101/2021.02.23.21252287

**Authors:** Semra Tibebu, Kevin A. Brown, Nick Daneman, Lauren A. Paul, Sarah A. Buchan

## Abstract

In this population-wide study in Ontario, Canada, we investigated the household secondary attack rate (SAR) to understand its relationship to household size and index case characteristics. We identified all patients with confirmed COVID-19 between July 1 and November 30, 2020. Cases within households were matched based on reported residential address; households were grouped based on the number of household contacts. The majority of households (68.2%) had a SAR of 0%, while 3,442 (11.7%) households had a SAR ≥75%. Overall household SAR was 19.5% and was similar across household sizes, but varied across index case characteristics. Households where index cases had longer delays between symptom onset and test seeking, households with older index cases, households with symptomatic index cases, and larger households located in diverse neighborhoods, were associated with greater household SAR. Our findings present characteristics associated with greater household SARs and proposes immediate testing as a method to reduce household transmission and incidence of COVID-19.

## Introduction

Household secondary attack rate (SAR) is an important indicator of the transmission of COVID-19. Previous studies have reported household SARs of COVID-19 ranging from 4% to 55%^1^. A majority of these studies were based on small cohort sizes, with few stratifying by number of household contacts^2^. In this population-wide study in Ontario, Canada, we investigated the household SAR to understand its relationship to household size and index case characteristics.

## Methods

We identified all patients with confirmed COVID-19 in Ontario’s provincial reportable disease surveillance system between July 1 and November 30, 2020. Cases within households were matched based on reported residential address^3^ and the number of household contacts were determined using reported household size. Cases living alone were excluded, as were those living in congregate settings such as retirement homes and shelters. Households were grouped based on household size.

We defined index cases based on the symptom onset date in the household; specimen collection date was used when symptom onset date was missing. In households with 1 contact, secondary cases were defined as those with symptom onset dates 1-14 days after the index case; in households with 2 or more contacts, this interval was increased to 1-28 days to allow for chains of transmission. Households containing multiple cases with the same earliest symptom onset date were excluded. In sensitivity analyses, secondary cases were those with onset 2-14 days after the index case. We obtained ethics approval from Public Health Ontario’s Research Ethics Board.

## Results

In this study period 68.5% of cases reported a household size. From those, we identified 29,352 households with 84,125 household contacts and 16,404 secondary cases. The majority of households (68.2%) had a SAR of 0% (Figure 1), while 3,442 (11.7%) households had a SAR ≥75%. Overall household SAR was 19.5% and was similar across household sizes, but varied across index case characteristics (Table 1). Households with index cases aged 20-30 years had the lowest SAR (16.5%) compared to other age groups. Households with symptomatic index cases had greater SAR compared to households with asymptomatic index cases (22.1% vs 6.2%); this was consistent across household sizes. Larger households in the most ethnically diverse neighborhoods had greater SAR than those in the least ethnically diverse neighborhoods (15.0% vs 20.3%); this was not evident in smaller households. Index cases with longer delays between symptom onset and test seeking were associated with greater household SAR; each one-day increase in testing delay was associated with a 1.8% increase in SAR. When secondary case onset was 2-14 days after the index case, household SAR was 15.5%.

**Table 1.**
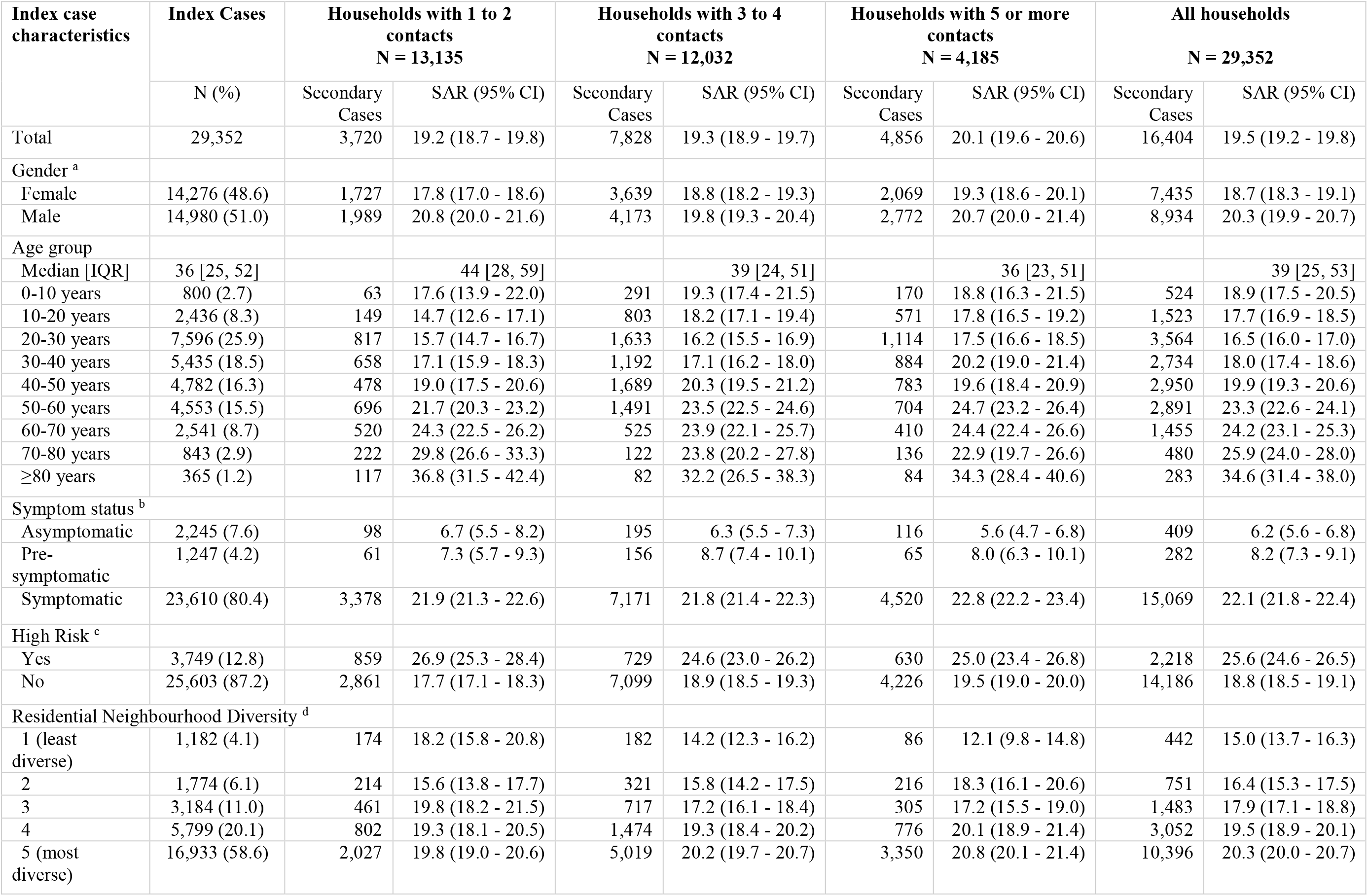
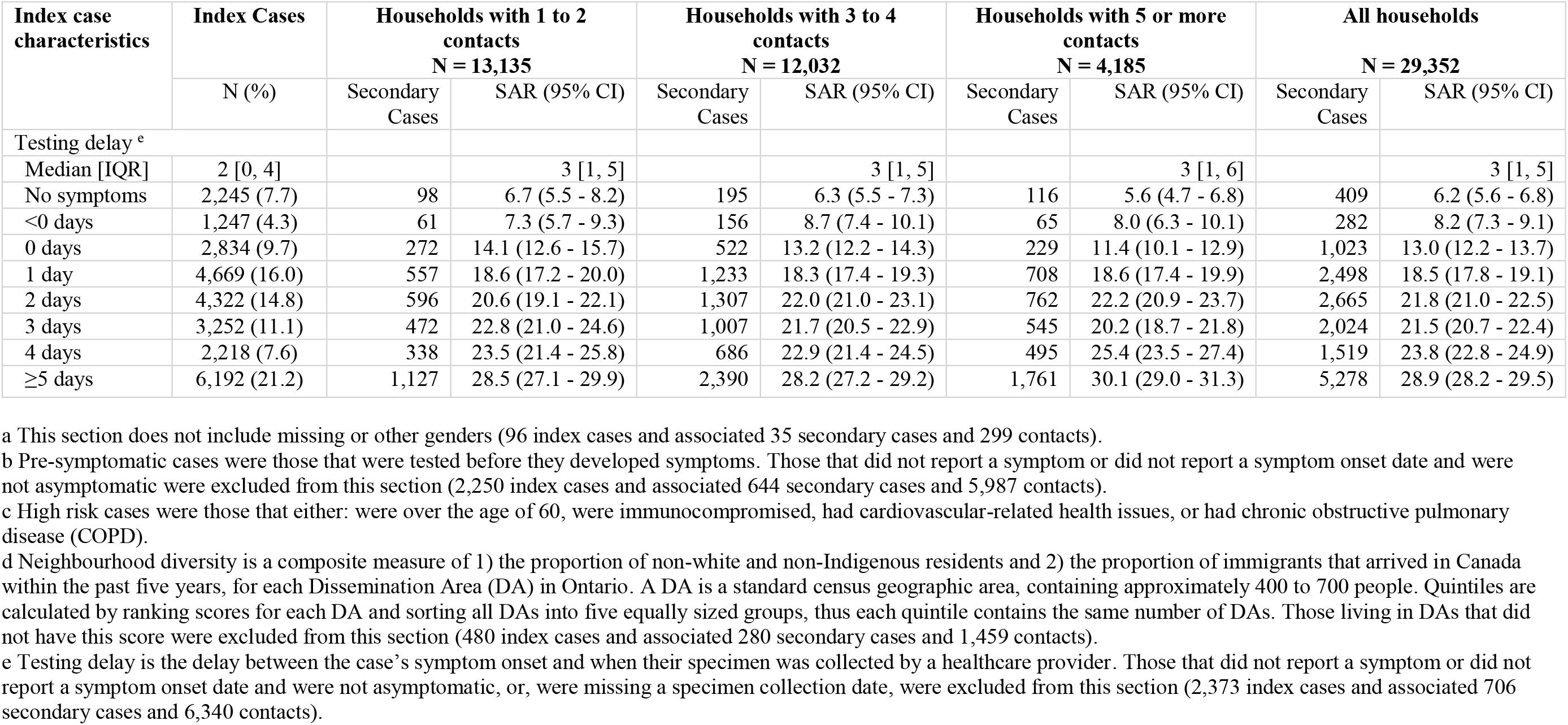
Number of secondary cases and household secondary attack rate by index case characteristics and the number of household contacts.

**Figure 1.**
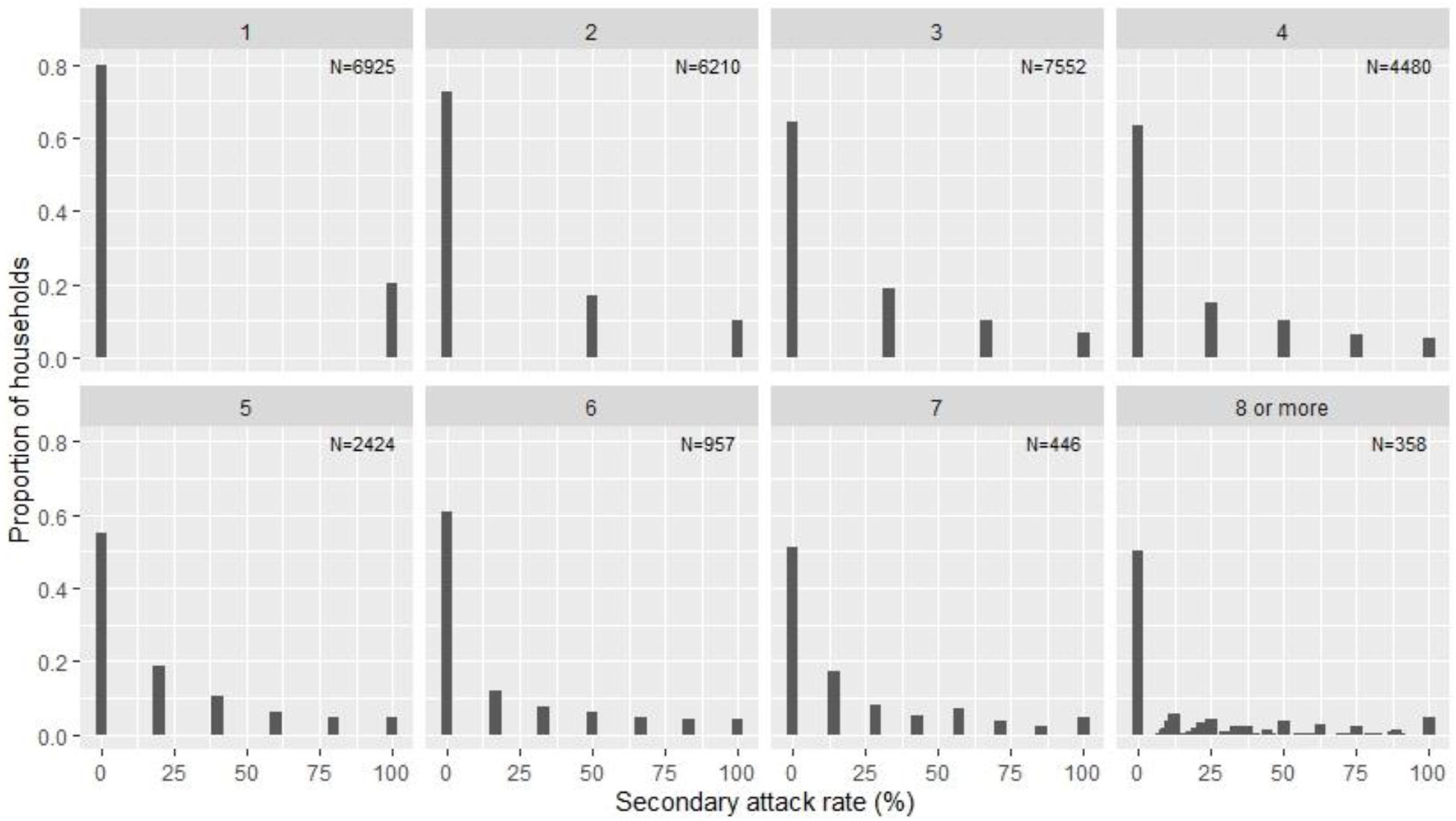
Distribution of household secondary attack rate (%) stratified by number of households contacts. a. N is the total number of households with the specified number of contacts.

## Discussion

Household SARs differed by index case characteristics. Our estimated household SAR (19.5%) aligns with a pooled estimate compiled by Koh et al. (18.1%, 95% CI: 15.7-20.6). Unlike our findings, studies have reported greater SARs in households with more contacts, though these studies utilized cohorts with fewer than 1000 participants and conducted enhanced contact follow-up^2,4^. Asymptomatic index cases associated with lower SAR is consistent with previous findings^1,2^.The increased SAR within larger households in more ethnically diverse neighborhoods reflects findings that visible minorities are less likely to work from home during the pandemic^5^ and that greater household crowding in these communities contribute to their increased risk of COVID-19^6^. Our findings propose the importance of immediate test seeking, as a confirmed case may motivate behavior to prevent household transmission. This analysis is limited by the absence of household contact information (including COVID-19 testing status), and by the potential misclassification of index cases, especially in households with asymptomatic cases. Our findings present characteristics associated with greater household SARs and proposes immediate testing as a method to reduce household transmission and incidence of COVID-19.

## Data Availability

The authors had full access to all data in the study and accept responsibility for the decision to submit for publication.

